# A nationwide deep learning pipeline to predict stroke and COVID-19 death in atrial fibrillation

**DOI:** 10.1101/2021.12.20.21268113

**Authors:** Alex Handy, Angela Wood, Cathie Sudlow, Christopher Tomlinson, Frank Kee, Johan H Thygesen, Mohammad Mamouei, Reecha Sofat, Richard Dobson, Samantha Ip, Spiros Denaxas, on behalf of the CVD-COVID-UK Consortium

## Abstract

Deep learning (DL) and machine learning (ML) models trained on long-term patient trajectories held as medical codes in electronic health records (EHR) have the potential to improve disease prediction. Anticoagulant prescribing decisions in atrial fibrillation (AF) offer a use case where the benchmark stroke risk prediction tool (CHA_2_DS_2_-VASc) could be meaningfully improved by including more information from a patient’s medical history. In this study, we design and build the first DL and ML pipeline that uses the routinely updated, linked EHR data for 56 million people in England accessed via NHS Digital to predict first ischaemic stroke in people with AF, and as a secondary outcome, COVID-19 death. Our pipeline improves first stroke prediction in AF by 17% compared to CHA_2_DS_2_-VASc (0.61 (0.57-0.65) vs 0.52 (0.52-0.52) area under the receiver operating characteristics curves, 95% confidence interval) and provides a generalisable, opensource framework that other researchers and developers can build on.

## MAIN

Recent advances in artificial intelligence can provide the basis for improving medical predictions^1^. In particular, advances in modelling large sequences of text using deep learning (DL) and natural language processing^2,3^ has opened up the possibility of harnessing long-term patient trajectories held as medical codes in electronic health records (EHR)^4,5^. Unlike conventional statistical and machine learning (ML) models, DL models can learn representations by directly taking long, individual sequences of medical codes stored in EHRs as inputs and could potentially identify complex, long-term dependencies between medical events^6^. To date, the improved performance of these DL models on their selected prediction tasks is promising^4,5^ but there has been limited comparison against prediction tools used routinely in clinical practice with comparisons typically made to other DL or ML methods. A direct comparison is important to demonstrate clearly where and by how much DL and ML can offer improvements and to help in integrating these methods (where appropriate) into routine clinical practice.

Anticoagulant prescribing decisions in atrial fibrillation (AF) offer a use case where the benchmark stroke risk prediction tool (CHA_2_DS_2_-VASc^7^) used routinely in clinical practice could be meaningfully improved by including more information from a patient’s medical history. AF is a disturbance of heart rhythm affecting 37.5 million people globally^8^ and significantly increases ischaemic stroke risk^9^. Anticoagulants reduce the risk of stroke^10^ and are recommended for people with AF and a high risk of stroke, broadly defined as a CHA_2_DS_2_-VASc >=2 based on the National Institute for Health and Care Excellence (NICE) threshold^11,12^. The CHA_2_DS_2_-VASc score benefits from being easy to calculate and interpret, however, it only measures 7 variables (age, sex, history of congestive heart failure, hypertension, stroke/TIA/thromboembolism, vascular disease and diabetes) and NICE’s own evidence review highlights the need for improved stroke risk assessment^13^. It shows that whilst CHA_2_DS_2_-VASc is good for identifying people potentially at risk of stroke (high sensitivity) it is poor at identifying people who may not have a stroke (low specificity)^14^. The ability of CHA_2_DS_2_-VASc to discriminate an individual’s future stroke risk is also only moderate (pooled area under the receiver operating characteristics curve (AUC) of 0.67 across 27 studies^14^) and potentially lower for predicting first ever stroke based on information at the point of AF diagnosis where available evidence is significantly limited. Recent research has also observed that pre-existing use of antithrombotics, particularly anticoagulants, is associated with lower odds of people with AF dying from COVID-19^15,16^. A model that could improve prediction of first stroke in people with AF and also identify those at greatest risk of COVID-19 death would be a potentially valuable new tool to inform anticoagulant prescribing decisions.

In this study, we design and build the first DL and ML pipeline that uses the routinely updated, linked EHR data for 56 million people in England accessed via NHS Digital’s Trusted Research Environment (TRE)^17^. We use this pipeline to predict first ischaemic stroke in people with AF (mean follow-up time 7.2 years), and as a secondary outcome, COVID-19 death, using individual sequences of medical codes from the entire primary and secondary care record.

We compare the performance of our DL and ML pipeline directly against the CHA_2_DS_2_-VASc score to support translation to clinical practice and demonstrate a 17% improvement on predicting first stroke in AF.

The code for our pipeline is generalisable, opensource and designed to provide a proof-of-concept framework that other researchers and developers can build on.

## RESULTS

### Nationwide deep learning and machine learning pipeline

We built our DL and ML pipeline within NHS Digital’s TRE for England which provides secure, remote access to routinely collected, linked, person level EHR data for over 56 million people^17^. Available data sources include primary care, secondary care, pharmacy dispensing, death registrations and COVID-19 tests and vaccines.

For this study, we constructed individual sequences of medical codes using all coded events from the General Practice Extraction Service Extract for Pandemic Planning and Research (GDPPR) and Hospital Episode Statistics on admissions (HES APC – primary diagnosis code) datasets. Recorded events were organised into a time ordered list (earliest first) of medical codes (e.g. [SNOMED-CT code 1, ICD-10 code 1, SNOMED-CT code 2, ICD-10 code 2…SNOMED-CT n / ICD-10 code n]) up to the target inclusion event (e.g. first AF diagnosis) alongside a set of static variables that represent demographic information (e.g. female, age at first AF diagnosis, ethnicity, see *Figure 1*).

**Figure 1.**
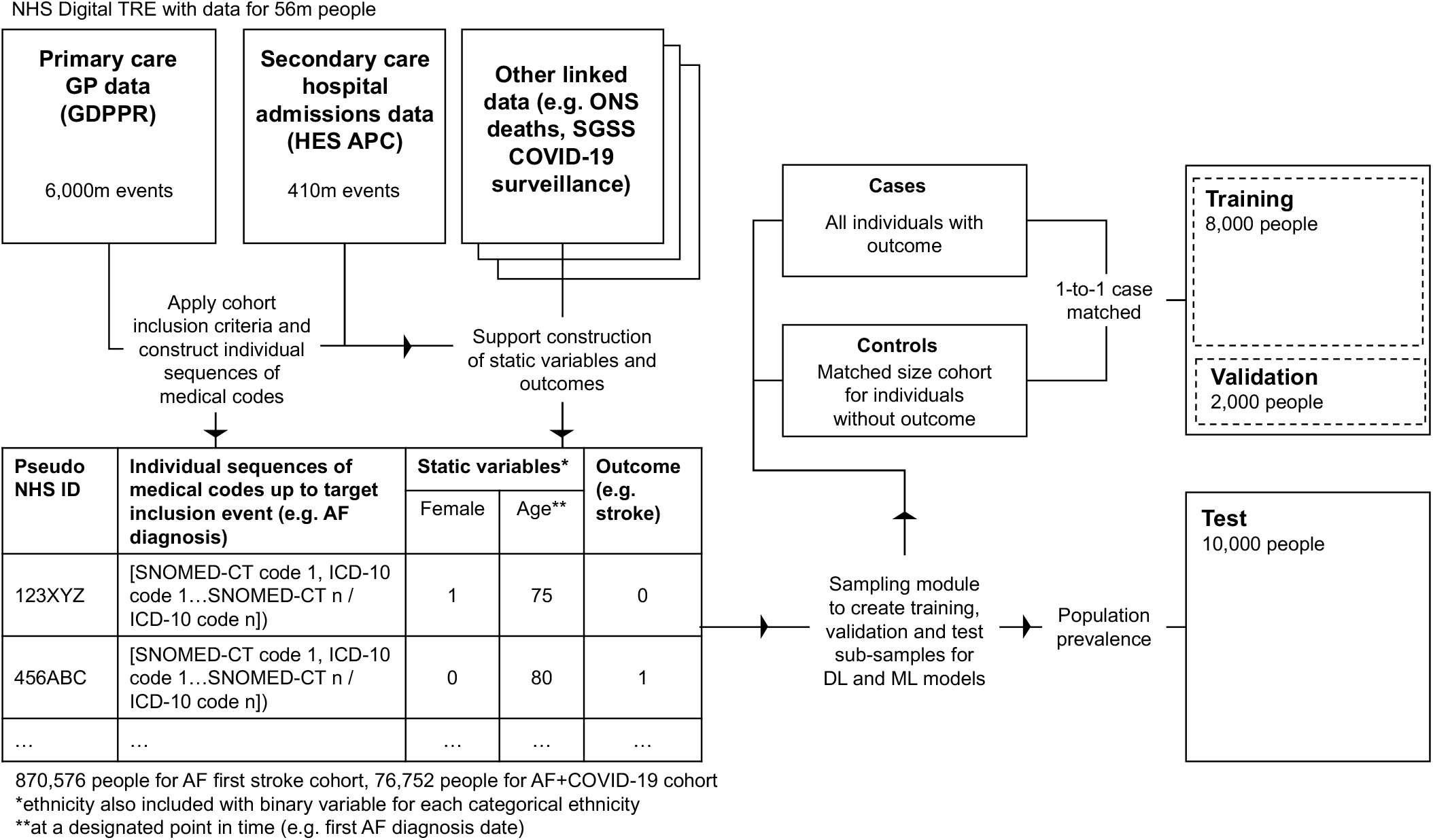
overview of the key data processing steps for the DL and ML pipeline within NHS Digital’s TRE for England

To make the analysis computationally tractable, we built a sampling module which creates training, validation and test sub-samples (with cohort inclusion criteria applied) from this transformed data.

For the model components, transformer and long short-term memory (LSTM) network architectures were selected as blueprints for the DL models due to their suitability for sequence modelling both within and outside of EHRs^3–5,18–20^. For the ML models, logistic regression, random forest and XGBoost were selected to provide a conventional benchmark (logistic regression) and a selection of models with evidence of performing well on structured, tabular data (random forest and XGboost)^21,22^. CHA_2_DS_2_-VASc scores were also calculated for each individual (see *“Methods”* section) with CHA_2_DS_2_-VASc >= 2 used as the baseline for assessing the prediction tasks.

For the ML models (logistic regression, random forest and XGBoost) individual sequences of medical codes were represented as one hot encoded variables for each unique code in the cohort sample with static variables represented as covariates in their continuous or categorical form. The DL models (transformer and LSTM) required a more sophisticated input representation and architecture (see *Figure 2* and “*Methods”* section) that preserved the sequential order of medical codes for each individual. The same model architectures were used for all prediction tasks.

**Figure 2.**
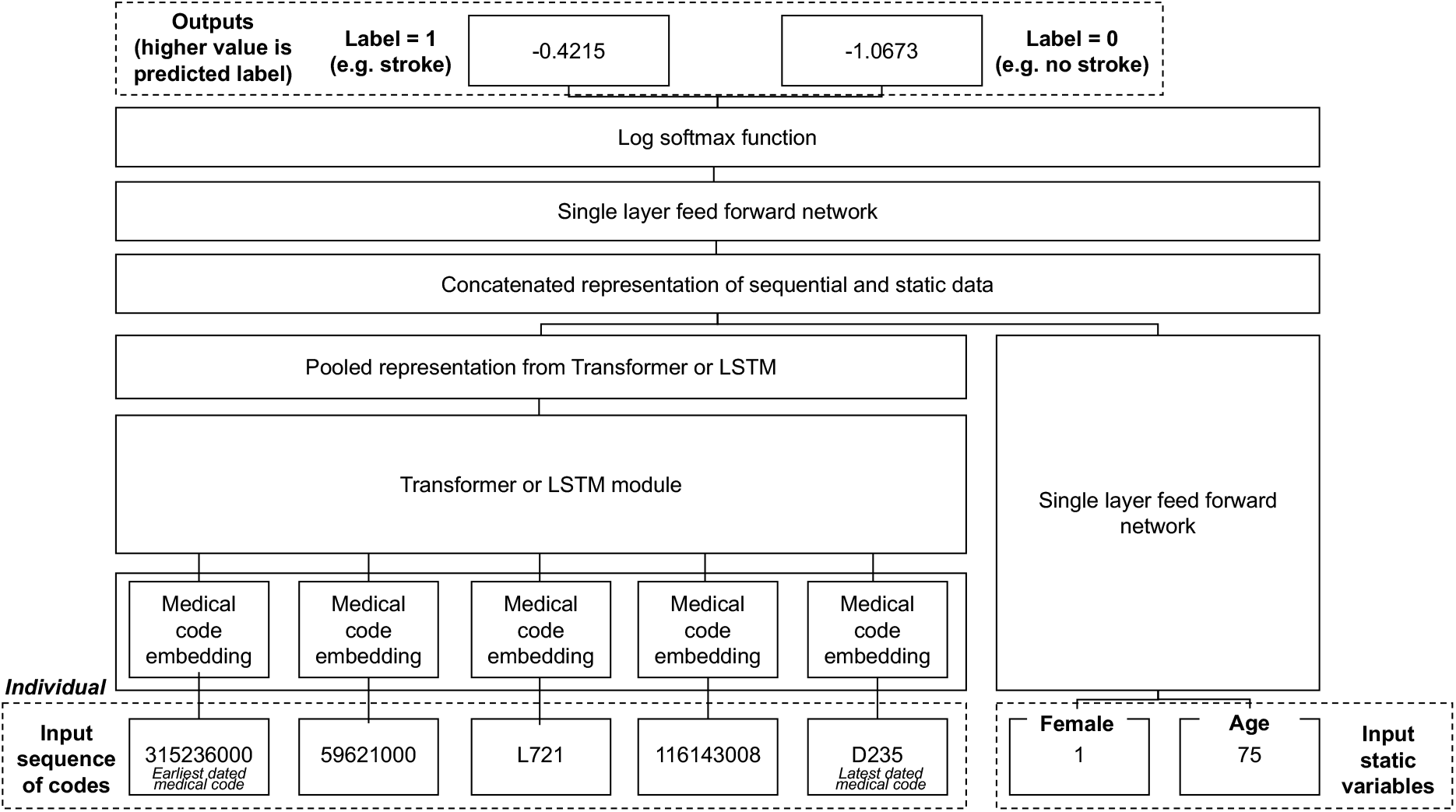
overview of the input representation and architecture for the transformer and LSTM DL models. *Note, length and number of input variables are illustrative, for precise specifications see “Methods” section*.

### Cohort characteristics for prediction tasks

Two sample cohorts were created for the prediction tasks, an AF first stroke cohort (an AF diagnosis and no prior stroke diagnosis) and an AF+COVID-19 cohort (an AF diagnosis, no prior stroke diagnosis and a positive COVID-19 event, see *“Methods”* section for more details).

From a total of 55,903,113 people registered with a GP practice in England and alive on January 1^st^ 2020, 870,576 had a diagnosis of AF in their GP record (and met the other inclusion criteria) and 16,563 (1.9%) had a first ischaemic stroke after their AF diagnosis up to May 1^st^ 2021.

The AF+COVID-19 cohort contained 76,752 people of whom 17,822 (23.2%) died of COVID-19. A flow chart with inclusion criteria is included in *Supplementary Figure 1* and a summary of the demographic and medical characteristics of both cohorts is included in *Table 1*.

**Table 1.** summary of demographic and medical characteristics for first stroke and COVID-19 cohort *\* The number of people with COVID-19 events is lower in first stroke cohort due to the date of first AF diagnosis cut-off being several years earlier (on average) than the first COVID-19 event date cut-off and as a result a larger proportion of people did not have >=5 recorded medical codes prior to this date (see “Methods” section).*

|  | **AF first stroke cohort** | **AF+COVID-19 cohort** |
| --- | --- | --- |
| Individuals | 870576 | 76752 |
| Age (mean years, +/- sd) |  |  |
| Age at Jan 1st 2020 | 76 (+/- 11.8) | 77 (+/- 13.6) |
| Age at first AF diagnosis | 69 (+/- 12.3) | 69 (+/- 14.6) |
| Follow-up time post AF diagnosis (mean years, +/- sd) | 7.2 (+/- 4.9) | 8.7 (+/- 6.7) |
| Female | 376398 (43.2%) | 33664 (43.9%) |
| Ethnicity |  |  |
| White | 832291 (95.7%) | 71794 (93.5%) |
| Asian or asian british | 18823 (2.2%) | 2794 (3.6%) |
| Black or black british | 8781 (1.0%) | 1044 (1.4%) |
| Mixed ethnicity | 3131 (0.4%) | 363 (0.5%) |
| Other ethnic groups | 6920 (0.8%) | 756 (1.0%) |
| Medical characteristics |  |  |
| All recorded medical codes (mean count up to target inclusion event, +/- sd) | 71 (+/- 86.5) | 418 (+/- 319.7) |
| Unique recorded medical codes (mean count up to target inclusion event, +/- sd) | 20 (+/- 16.5) | 76 (+/- 27.6) |
| Ischaemic stroke | 16563 (1.9%) | 2524 (3.3%) |
| COVID-19 event* | 67208 (7.7%) | 76730 (100%) |
| COVID-19 death | 15899 (1.8%) | 17822 (23.2%) |

### Prediction task performance

The primary prediction task was to predict the binary outcome of first ischaemic stroke in people with AF (mean follow-up time 7.2 years). For this task, XGBoost was the top performing model (AUC=0.61 (0.57-0.65) with random forest a close second (AUC=0.60 (0.58-0.62), followed by transformer (AUC=0.58 (0.58-0.58) and logistic regression (AUC=0.58 (0.56-0.60) models. LSTM (AUC=0.53 (0.47-0.59) and CHA_2_DS_2_-VASc >=2 (AUC=0.52 (0.52-0.52) were the worst performing models (see *Table 2*). XGBoost did not have the highest overall accuracy (0.53 (0.51-0.55) vs random forest with 0.56 (0.52-0.60) but did have higher sensitivity (0.68 (0.62-0.74) vs 0.65 (0.63-0.67)).

**Table 2.**
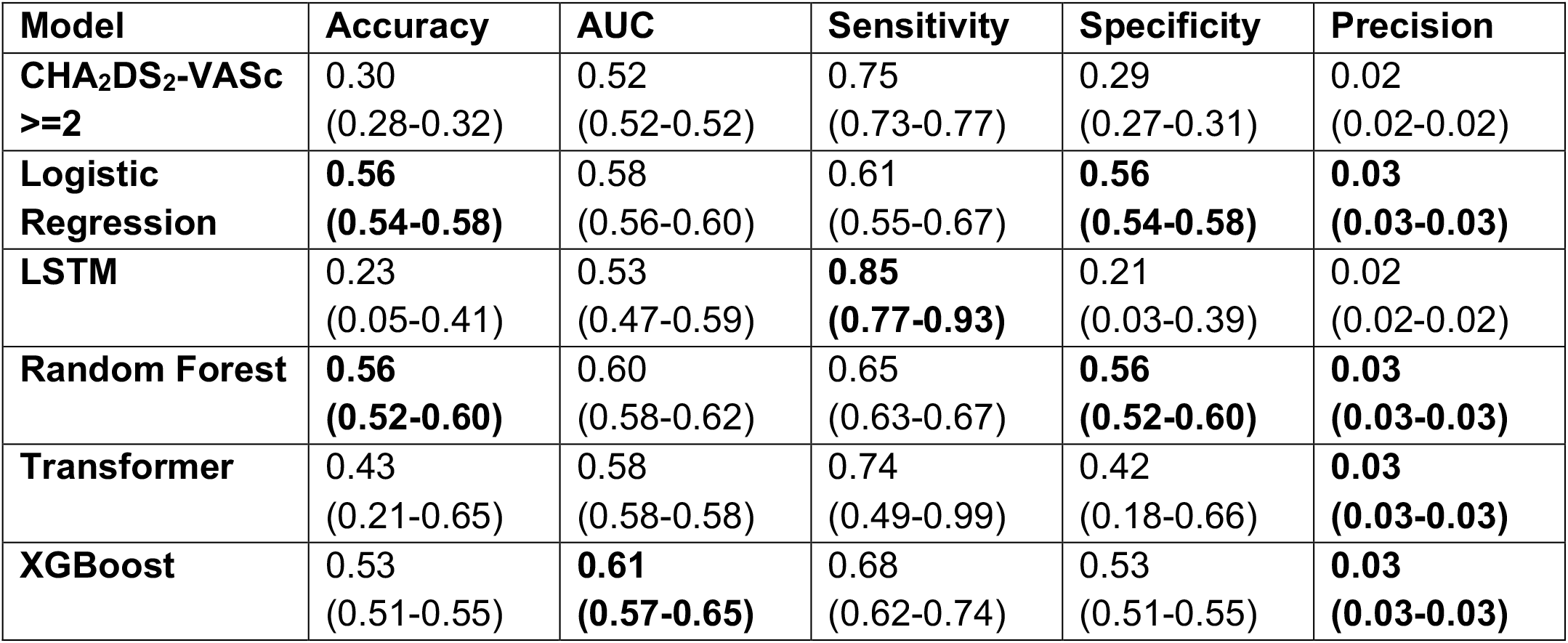
summary of model performance statistics for predicting first ischaemic stroke after AF in all groups. 95% confidence intervals (CI) in brackets. Bold font marks highest performing model for each metric.

| Model | Accuracy | AUC | Sensitivity | Specificity | Precision |
| --- | --- | --- | --- | --- | --- |
| **CHA<sub>2</sub>DS<sub>2</sub>-VASc**<br>**>=2** | 0.30<br>(0.28-0.32) | 0.52<br>(0.52-0.52) | 0.75<br>(0.73-0.77) | 0.29<br>(0.27-0.31) | 0.02<br>(0.02-0.02) |
| **Logistic Regression** | **0.56**<br>**(0.54-0.58)** | 0.58<br>(0.56-0.60) | 0.61<br>(0.55-0.67) | **0.56**<br>**(0.54-0.58)** | **0.03**<br>**(0.03-0.03)** |
| **LSTM** | 0.23<br>(0.05-0.41) | 0.53<br>(0.47-0.59) | **0.85**<br>**(0.77-0.93)** | 0.21<br>(0.03-0.39) | 0.02<br>(0.02-0.02) |
| **Random Forest** | **0.56**<br>**(0.52-0.60)** | 0.60<br>(0.58-0.62) | 0.65<br>(0.63-0.67) | **0.56**<br>**(0.52-0.60)** | **0.03**<br>**(0.03-0.03)** |
| **Transformer** | 0.43<br>(0.21-0.65) | 0.58<br>(0.58-0.58) | 0.74<br>(0.49-0.99) | 0.42<br>(0.18-0.66) | **0.03**<br>**(0.03-0.03)** |
| **XGBoost** | 0.53<br>(0.51-0.55) | **0.61**<br>**(0.57-0.65)** | 0.68<br>(0.62-0.74) | 0.53<br>(0.51-0.55) | **0.03**<br>**(0.03-0.03)** |

Models were also evaluated on sub-groups to compare predictive performance across gender, age and ethnicities (see *Supplementary Table 1*). Performance was broadly consistent across sub-groups with the exception of potentially lower performance in individuals with a recorded ethnicity of “black or black british” (XGBoost AUC=0.30 (0.00-0.93)), “mixed ethnicity” (XGBoost AUC=0.33 (0.00-1.00)) or “other ethnic group” (XGBoost AUC=0.49 (0.00-1.00)).

For our secondary outcome, COVID-19 death, XGBoost was also the top performing model (AUC=0.73 (0.73-0.73), (see *Table 3)* followed by random forest (AUC=0.70 (0.68-0.72, transformer (AUC=0.69 (0.67-0.71)), logistic regression (AUC=0.69 (0.67-0.71) and the LSTM model (AUC=0.67 (0.65-0.69). CHA_2_DS_2_-VASc >=2 (AUC=0.58 (0.58-0.58) was the worst performing model by a greater distance than in the stroke prediction task. XGBoost was also the top performing model for accuracy (0.69 (0.69-0.69) and precision (0.42 (0.42-0.42)) but had marginally lower sensitivity than random forest (0.81 (0.77-0.85) and CHA_2_DS_2_-VASc >=2 (0.98 (0.98-0.98), which labelled nearly all COVID-19 deaths.

**Table 3.** summary of model performance statistics for predicting COVID-19 death in people diagnosed with AF (and no prior stroke diagnosis) in all groups. 95% CI in brackets. Bold font marks highest performing model for each metric.

| Model | Accuracy | AUC | Sensitivity | Specificity | Precision |
| --- | --- | --- | --- | --- | --- |
| **CHA<sub>2</sub>DS<sub>2</sub>-VASc**<br>**>=2** | 0.37<br>(0.37-0.37) | 0.58<br>(0.58-0.58) | **0.98**<br>**(0.98-0.98)** | 0.19<br>(0.19-0.19) | 0.27<br>(0.27-0.27) |
| **Logistic Regression** | 0.68<br>(0.68-0.68) | 0.69<br>(0.67-0.71) | 0.70<br>(0.66-0.74) | **0.68**<br>**(0.68-0.68)** | 0.40<br>(0.40-0.40) |
| **LSTM** | 0.63<br>(0.49-0.77) | 0.67<br>(0.65-0.69) | 0.75<br>(0.53-0.97) | 0.59<br>(0.34-0.84) | 0.36<br>(0.28-0.44) |
| **Random Forest** | 0.65<br>(0.65-0.65) | 0.70<br>(0.68-0.72) | 0.81<br>(0.77-0.85) | 0.60<br>(0.60-0.60) | 0.38<br>(0.38-0.38) |
| **Transformer** | 0.67<br>(0.61-0.73) | 0.69<br>(0.67-0.71) | 0.72<br>(0.66-0.78) | 0.66<br>(0.56-0.76) | 0.39<br>(0.35-0.43) |
| **XGBoost** | **0.69**<br>**(0.69-0.69)** | **0.73**<br>**(0.73-0.73)** | 0.79<br>(0.77-0.81) | 0.66<br>(0.66-0.66) | **0.42**<br>**(0.42-0.42)** |

In contrast to the stroke prediction task, there was more divergence in model performance across sub-groups (see *Supplementary Table 2*). AUC was 4% (XGBoost) to 13% (CHA_2_DS_2_-VASc) lower for women compared to men. For people aged under 65, CHA_2_DS_2_-VASc, AUC was 33% higher than for people aged 65 or over. However, in the other models AUC was 23% (XGBoost) to 5% (Transformer / LSTM) lower for people aged under 65 compared to people aged 65 or over. Across ethnicities, performance was more consistent, with the exception of people with a “mixed” recorded ethnicity where AUC was on average 11% higher than for the other ethnic groups.

## DISCUSSION

This study is the first to design and build a DL and ML pipeline that uses the routinely updated, linked EHR data for 56 million people in England accessed via NHS Digital. All the DL and ML models outperformed CHA_2_DS_2_-VASc for predicting first ischaemic stroke (AUC=0.52 (0.52-0.52)) and COVID-19 death (AUC=0.58 (0.58-0.58)) in people with AF. However, DL models did not outperform more conventional ML methods, with XGBoost the top performing model for predicting first stroke (AUC=0.61 (0.57-0.65)) and COVID-19 death (AUC=0.73 (0.73-0.73). We also provide detailed performance statistics (e.g. accuracy, sensitivity, specificity, precision) and sub-group analysis (e.g. gender, age, ethnicity) to ensure we adhere to recommended guidance for reporting on DL and ML analysis in clinical research^23,24^ wherever possible and appropriate. Whilst the top performing model improves first stroke prediction by 17% compared to CHA_2_DS_2_-VASc (0.61 vs 0.52), further improvements are required before our pipeline could be considered for use in clinical practice.

Despite the rapid improvements in DL and ML, applications in routine clinical care remain challenging^25^. This is partly because DL and ML applications are developed on data and infrastructure which is often different to what is available in routine care^25^. NHS Digital’s TRE for England is unique in providing a platform with widely linked, routinely collected, population-scale data that could form the foundation of a nationwide DL and ML powered EHR^17^. Motivated by this potential we have successfully developed the first proof-of-concept DL and ML pipeline that deploys advanced DL and ML models within the unique environment of NHS Digital’s TRE for England. We then sought to demonstrate that this could improve a clinical use case where a prediction tool is already routinely used and where there is an opportunity to improve performance by harnessing high-dimensional information from a patient’s medical history. We, therefore, selected anticoagulant prescribing decisions in AF and tested our pipeline against the CHA_2_DS_2_-VASc score for predicting first stroke after AF and COVID-19 death.

It is already recognised that CHA_2_DS_2_-VASc is an imprecise tool for stroke prediction^14^ and this is reflected in the differing thresholds recommended in international clinical guidance^13,26^. Our findings reinforce the challenges of precisely predicting stroke using CHA_2_DS_2_-VASc, particularly for predicting first ever stroke at the point of AF diagnosis, where the performance on discriminating between someone who had a stroke compared to someone who did not was little better than chance (an AUC of 0.50) in our study. The poor performance of CHA_2_DS_2_-VASc may be partly explained by its heavy weighting of previous stroke diagnoses which get 2 points in a 7 variable, 9 point system. Our DL and ML models improved prediction performance, potentially by being able to use more variables (represented as medical codes) from an individual’s primary and secondary care record up to AF diagnosis. However, the sequential ordering of an individual’s medical codes did not appear to improve predictions as XGBoost outperformed the DL models using only the binary information of whether a person had a recorded medical code (one-hot encoding). XGBoost has been shown to perform as well as DL models on tabular data^22^ and has also been applied to a range of EHR disease prediction problems^27,28^. Despite a 17% improvement compared to CHA_2_DS_2_-VASc, the performance was still only moderate and supports the observation that predicting first stroke in atrial fibrillation is a challenging prediction problem.

The performance on prediction of COVID-19 death was more encouraging and begins to demonstrate how our DL and ML pipeline could deliver larger improvements to disease prediction and be used in clinical practice. A key driver of the improved performance of predicting COVID-19 death is likely the inclusion of more medical codes due to the later, on average, target inclusion event of a positive COVID-19 event compared to a first AF diagnosis. This meant that DL and ML models had access to, on average, 76 unique medical codes from an individual’s medical history vs 20 in the first stroke cohort. Importantly, all the models except CHA_2_DS_2_-VASc showed an AUC improvement of at least 17% (vs 12%) indicating that DL and ML architectures can extract incrementally valuable information from longer sequences.

There are also several key limitations which prevented us from maximising the potential performance from DL and ML architectures. Firstly, graphical processing units (GPUs) and some parallel computing methods are currently restricted on the NHS Digital’s TRE for England meaning that it was not possible to train models on larger datasets (e.g. 10,000+) or create DL architectures with more layers. This also prevented us from including an individual’s full medical history (e.g. no repeating medical codes) and only allowed us to include the 100 most recent medical codes up to the target inclusion event.

Secondly, the NHS Digital TRE for England does not yet facilitate the use or creation of code embeddings pre-trained with other models. This transfer learning approach was adopted by the teams behind BEHRT^4^ and MedBERT^5^ and builds on the performance gains demonstrated by large language models such as BERT^2^ and GPT-3^29^.

Lastly, medical codes stored in structured EHR data are just one type of data modality and do not reflect the full diversity of an individual’s medical history. Even before adding new types of data to the TRE such as genetics, imaging and free text, there are observational values such as systolic blood pressure and cholesterol / HDL ratio which could be included in future models.

In addition to addressing the above, the next phase of our work will aim to improve the clinical interpretability of our DL and ML pipeline. For this study, we chose to compare model performance primarily using AUC on binary outcomes (e.g. 1 for stroke, 0 for no stroke) to support direct comparison to CHA_2_DS_2_-VASc but our pipeline is capable of producing estimated probabilities which could be used by clinicians as confidence measures and by future researchers to assess model calibrations. We will also explore adding feature assessment mechanisms (e.g. attention visualisation^30^) to ML and DL models but recognise that this alone still falls short of the interpretability clinicians need^31^. It will also be important for future work to consider the implications of how missing data within the patients’ sequence of medical codes might affect the accuracy of predictions in subgroups (e.g. deprived versus affluent) and adapt the algorithms accordingly to ensure equity^32^. Lastly, we will explore the potential to integrate adapted survival analysis models^33^ which could allow more precise censoring of individuals and enhanced interpretability through mapping “nodes” to biological features.

In conclusion, we designed and built the first DL and ML pipeline that uses the routinely updated, linked EHR data for 56 million people in England and improved first stroke prediction by 17% compared to CHA_2_DS_2_-VASc. Further potential improvements could be achieved by using higher computation training regimes, pre-trained embeddings and more data modalities.

## METHODS

### Data sources

The DL and ML pipeline was created using NHS Digital’s TRE for England which provides secure, remote access to linked, person level EHR data for over 56 million people^17^. Available data sources include primary care, secondary care, pharmacy dispensing, death registrations and COVID-19 tests and vaccines. For this study, we constructed individual sequences of medical codes using all coded events from the General Practice Extraction Service Extract for Pandemic Planning and Research (GDPPR) and Hospital Episode Statistics on admissions (HES APC) datasets. AF and stroke diagnoses were determined from GDPPR and COVID-19 events and deaths from a combination of HES, COVID-19 Hospitalisations in England Surveillance System (CHESS), Public Health England’s Second Generation Surveillance System (SGSS), Secondary Uses Service (SUS) and Office for National Statistics (ONS) Civil Registration of Deaths.

### Cohort selection

Individuals were eligible for the sample cohorts if they had five or more recorded medical codes [as in BEHRT^4^] across GDPPR and HES APC, were >= 18 years old and alive on January 1st 2020, had available sex, ethnicity and GP practice location data (based on most recent, available data across primary care (GDPPR), secondary care (HES APC) and death registrations (Office for National Statistics)) and had a diagnosis of AF (coded in GDPPR). For the AF first stroke cohort, people who had a stroke diagnosis (including non-ischaemic strokes) prior to their AF diagnosis were excluded (see *Supplementary Figure 1* for a cohort inclusion flowchart). People with AF who had an ischaemic stroke diagnosis after their AF diagnosis, were only included if their stroke occurred two or more months after the date of their first AF diagnosis to help screen out delayed coding of cases which may have occurred prior to AF diagnosis.

For the AF+COVID-19 cohort, in addition to an AF diagnosis, individuals required a recorded COVID-19 event defined as any of a positive test (polymerase chain reaction or lateral flow), a coded diagnosis in primary or secondary care or a COVID-19 diagnosis on a death certificate^34^. The COVID-19 death outcome included people with a COVID-19 diagnosis on their death certificate in any position, a registered death within 28 days of their first recorded COVID-19 event or a discharge destination denoting death after a COVID-19 hospitalisation^34^. Follow-up for both first ever stroke and COVID-19 death was conducted from date of first event (AF diagnosis or COVID-19 event) up to May 1^st^ 2021. Further details on phenotyping algorithms used are available on GitHub (https://github.com/BHFDSC/CCU004_02/tree/main/phenotypes).

The entire eligible study population was then randomly split 80:20 into training and test datasets. Prevalence of first ischaemic stroke after AF was low (1.9%) which means the target class (stroke) was highly imbalanced in the training data. To address this for the training data, we created a rebalanced sample by selecting all stroke cases and randomly selecting (with replacement) controls at a ratio of 1 control to 1 stroke case. This ratio was selected after initial experimentation which showed that DL and ML models had limited ability to discriminate (based on AUC) after being trained on ratios of 1-to-3 and population prevalence. The testing dataset was kept at the population prevalence. The same approach was adopted for COVID-19 death which had moderate prevalence (23.2%) in people with AF.

As outlined in *Figure 1*, a sampling module was developed to create computationally tractable sub-samples from the nationwide, eligible study population. Random sub-samples of 10,000 people were selected from the rebalanced training and testing datasets. The training sub-sample was then split into model training data (n=8000) and validation data (n=2000), with the model with the highest AUC on the validation data selected for testing. To assess the reliability of model predictions, three versions of each training and test sub-sample were created with averages and confidence intervals reported in results.

The maximum length of medical codes included for each individual was also adjusted to reduce computational requirements. Models were trained and tested with a limit of 100 medical codes which included all codes for 99% of the AF first stroke cohort and >75% of AF+COVID-19 cohort.

### Statistical analysis and model implementation

The primary prediction task was to predict the binary outcome of first ischaemic stroke in people with AF.

A CHA_2_DS_2_-VASc score >=2 was used as the baseline with individuals with a score of >=2 assigned a label of 1 (prediction of future stroke) and those <2 assigned a label of 0 (prediction of no future stroke). The CHA_2_DS_2_-VASc score was calculated for each individual in the cohort based on the scoring system outlined here^7^, with “Stroke/TIA/thromboembolism history” excluded due to the removal of individuals with these diagnoses from the cohort given the target prediction outcome was *first* stroke.

For the ML models (logistic regression, random forest and XGBoost) individual sequences of medical codes were represented as one hot encoded variables for each unique code in the cohort sample with the static variables (female, age at first AF diagnosis, ethnicity) represented as covariates in their continuous or categorical form.

The DL models (transformer and LSTM) required a more sophisticated input representation and architecture (see *Figure 2* for graphical overview). The general design principle was to keep the models as simplistic as possible for the proof-of-concept with default configurations used where possible (refer to Pytorch documentation - https://pytorch.org/docs/stable/torch.html) and additional layers and modules kept to a minimum.

A vocabulary of each unique medical code from all the individual sequences of medical codes from the cohort sample was assembled and used to create a trainable set of vector embeddings for each medical code. Individual sequences of medical codes are, therefore, input into DL models as sequences with dimensions D^m*n^ with *m* the max length of an individual sequence of medical codes in the cohort (limited to 100 in this study and padded with zeros for individuals with shorter sequences) and *n* the size of the medical code vector embeddings (200 in this study). The transformer also has positional embeddings to ensure it has the ability to learn information from the relative position of the medical codes^3^. These sequences are then passed through a sequential module (recurrent gated cells for LSTM and multi-headed attention layers for transformer) to provide a pooled representation of each sequence. Both the LSTM and transformer have two internal layers; two hidden layers for the LSTM and two encoder layers (with two attention heads) for the transformer.

Static variables were represented and input into the models as a vector of continuous values and passed through a separate feed forward layer with a rectified linear unit activation function prior to concatenation with the pooled outputs of the sequential module. The concatenated layer containing sequential and static information is then passed through another two feed forward layers to produce a vector the size of the number of output labels (two for a binary outcome) that is converted into logits for the loss function using the LogSoftmax.

Dropout layers (with a probability of dropout of 0.20) were included in the sequential module and the concatenated outputs to help prevent the model overfitting to the data.

Training parameters were kept consistent across both LSTM and transformer architectures with 10 epochs, a batch size of 64 and a learning rate of 0.001 (using an ADAM optimizer^35^). Negative log likelihood was used as the loss function. Plots of performance metrics across the training, validation and test datasets were visually inspected to confirm that 10 epochs was sufficient to reach convergence on the prediction tasks.

NHS Digital’s TRE for England runs on a Databricks cluster with Runtime 6.4 for Machine Learning and an i3.xlarge 30.5GB memory, 4 core worker. At the time of analysis, there were no GPUs available nor were Spark ML’s parallelized helper functions whitelisted for use. Data preparation, analysis and model building was performed using Python 3.7 and Spark SQL (2.4.5) with Databricks. The logistic regression and random forest models were built using the Python sklearn package (0.24.2) and fit with their default configurations (refer to sklearn documentation - https://scikit-learn.org/stable/modules/classes.html), with the exception of max iterations being set to 3000 for the logistic regression model. XGboost was built using the xgboost package (0.90) and fit with a “binary:logistic” objective and the remaining parameters as their default configuration. Both DL models were built with the PyTorch package (1.9.0). Accuracy, AUC, sensitivity, specificity and precision were estimated for each model using the sklearn. Summary tables were created using R version 4.0.3.

For any further specifications please refer to the code on GitHub (https://github.com/BHFDSC/CCU004_02/tree/main/code).

### Ethical and regulatory approvals

The data used in this study are available in NHS Digital’s TRE for England, but as restrictions apply they are not publicly available (https://digital.nhs.uk/coronavirus/coronavirus-data-services-updates/trusted-research-environment-service-for-england). The CVD-COVID-UK/COVID-IMPACT programme led by the BHF Data Science Centre (https://www.hdruk.ac.uk/helping-with-health-data/bhf-data-science-centre/) received approval to access data in NHS Digital’s TRE for England from the Independent Group Advising on the Release of Data (IGARD) (https://digital.nhs.uk/about-nhs-digital/corporate-information-and-documents/independent-group-advising-on-the-release-of-data) via an application made in the Data Access Request Service (DARS) Online system (ref. DARS-NIC-381078-Y9C5K) (https://digital.nhs.uk/services/data-access-request-service-dars/dars-products-and-services). The CVD-COVID-UK/COVID-IMPACT Approvals & Oversight Board (https://www.hdruk.ac.uk/projects/cvd-covid-uk-project/) subsequently granted approval to this project to access the data within the TRE for England. The de-identified data used in this study were made available to accredited researchers only. Analyses were conducted by approved researcher (AH) via secure remote access to the TRE. Only summarised, aggregate results were exported, following manual review by the NHS Digital ‘safe outputs’ escrow service, to ensure no output placed in the public domain contains information that may be used to identify an individual^17^. The North East-Newcastle and North Tyneside 2 research ethics committee provided ethical approval for the CVD-COVID-UK/COVID-IMPACT research programme (REC No 20/NE/0161).

### Patient and public involvement

The UK National Institute for Health Research-BHF Cardiovascular Partnership lay panel comprising individuals affected by cardiovascular disease reviewed and approved this project.

## Data Availability

The data used in this study are available in NHS Digital's TRE for England, but as restrictions apply they are not publicly available (https://digital.nhs.uk/coronavirus/coronavirus-data-services-updates/trusted-research-environment-service-for-england). Researchers can apply to the CVD-COVID-UK/COVID-IMPACT Approvals & Oversight Board (https://www.hdruk.ac.uk/projects/cvd-covid-uk-project/) for access.

## ACKNOWLEDGEMENTS

This study was carried out with the support of the BHF Data Science Centre led by HDR UK (BHF Grant no. SP/19/3/34678). This study makes use of de-identified data held in NHS Digital’s TRE for England and made available via the BHF Data Science Centre’s CVD-COVID-UK/COVID-IMPACT consortium. This study uses data provided by patients and collected by the NHS as part of their care and support. We would also like to acknowledge all data providers who make health relevant data available for research.

The views expressed are those of the authors and not necessarily those of the organisations listed. The funders of this work played no role in the collection, analysis, or interpretation of data; in the writing of the report; or in the decision to submit the article for publication.

## CONTRIBUTIONS

All authors drafted and reviewed the manuscript.

AH led the design and implementation of the analysis and is the guarantor.

CT, JHT, MM, SI and SD supported on the design and quality assurance of the data preparation and analysis code.

AW, CS, FK, RS, RD and SD supported on the overall study design and provided clinical expertise.

CS is the Director of the BHF Data Science Centre and coordinated approvals for and access to data within NHS Digital’s TRE for England for CVD-COVID-UK/COVID-IMPACT.

Members of the wider CVD-COVID-UK/COVID-IMPACT consortium (https://www.hdruk.ac.uk/wp-content/uploads/2021/12/211220-CVD-COVID-UK-COVID-IMPACT-Consortium-Members.pdf) also provided comments on drafts of the protocol and manuscript.

## COMPETING INTERESTS

The authors have no financial relationships with any organisations that might have an interest in the submitted work in the previous three years and no other relationships or activities that could appear to have influenced the submitted work.

## FUNDING

The British Heart Foundation Data Science Centre (grant No SP/19/3/34678, awarded to Health Data Research (HDR) UK) funded co-development (with NHS Digital) of the trusted research environment, provision of linked datasets, data access, user software licences, computational usage, and data management and wrangling support, with additional contributions from the HDR UK data and connectivity component of the UK governments’ chief scientific adviser’s national core studies programme to coordinate national covid-19 priority research. Consortium partner organisations funded the time of contributing data analysts, biostatisticians, epidemiologists, and clinicians.

AH is supported by research funding from the HDR UK text analytics implementation project.

AW is supported by the BHF-Turing Cardiovascular Data Science Award (BCDSA\100005) and by core funding from UK MRC (MR/L003120/1), BHF (RG/13/13/30194; RG/18/13/33946), and NIHR Cambridge Biomedical Research Centre (BRC-1215-20014).

CT is supported by a UCL UKRI Centre for Doctoral Training in AI-enabled Healthcare studentship (EP/S021612/1), MRC Clinical Top-Up and a studentship from the NIHR Biomedical Research Centre at University College London Hospital NHS Trust.

MM is supported by the Oxford Martin School (OMS), funded by the National Institute for Health Research (NIHR) Oxford Biomedical Research Centre (BRC), PEAK Urban programme, funded by the UKRI’s Global Challenge Research Fund Grant Ref: ES/P011055/1, and Novo Nordisk.

RD is supported by the following: (1) NIHR Biomedical Research Centre at South London and Maudsley NHS Foundation Trust and King’s College London, London, UK; (2) Health Data Research UK, which is funded by the UK Medical Research Council, Engineering and Physical Sciences Research Council, Economic and Social Research Council, Department of Health and Social Care (England), Chief Scientist Office of the Scottish Government Health and Social Care Directorates, Health and Social Care Research and Development Division (Welsh Government), Public Health Agency (Northern Ireland), British Heart Foundation and Wellcome Trust; (3) The BigData@Heart Consortium, funded by the Innovative Medicines Initiative-2 Joint Undertaking under grant agreement No. 116074. This Joint Undertaking receives support from the European Union’s Horizon 2020 research and innovation programme and EFPIA; it is chaired by DE Grobbee and SD Anker, partnering with 20 academic and industry partners and ESC; (4) the National Institute for Health Research University College London Hospitals Biomedical Research Centre; (5) the National Institute for Health Research (NIHR) Biomedical Research Centre at South London and Maudsley NHS Foundation Trust and King’s College London; (6) the UK Research and Innovation London Medical Imaging & Artificial Intelligence Centre for Value Based Healthcare; (7) the National Institute for Health Research (NIHR) Applied Research Collaboration South London (NIHR ARC South London) at King’s College Hospital NHS Foundation Trust.

SI is supported by the International Alliance for Cancer Early Detection, a partnership between Cancer Research UK C18081/A31373, Canary Center at Stanford University, the University of Cambridge, OHSU Knight Cancer Institute, University College London and the University of Manchester.

SD is supported by: (1) Health Data Research UK London, which receives its funding from HDR UK funded by the UK MRC, EPSRC, ESRC, Department of Health and Social Care (England), Chief Scientist Office of the Scottish Government Health and Social Care Directorates, Health and Social Care Research and Development Division (Welsh government), Public Health Agency (Northern Ireland), BHF, and Wellcome Trust; (2) The NIHR Biomedical Research Centre at University College London Hospital NHS Trust; (3) The Alan Turing Institute (EP/N510129/1); (4) The British Heart Foundation Accelerator Award (AA/18/6/24223); (5) The BigData@Heart Consortium, funded by the Innovative Medicines Initiative-2 Joint Undertaking under grant agreement No. 116074. This Joint Undertaking receives support from the European Union’s Horizon 2020 research and innovation programme and EFPIA; it is chaired by DE Grobbee and SD Anker, partnering with 20 academic and industry partners and ESC.

AW, RD and SD are part of the BigData@Heart Consortium, funded by the Innovative Medicines Initiative-2 Joint Undertaking under grant agreement No 116074.

## SUPPLEMENTARY MATERIAL

## SUPPLEMENTARY FIGURES

**Supplementary Figure 1.**
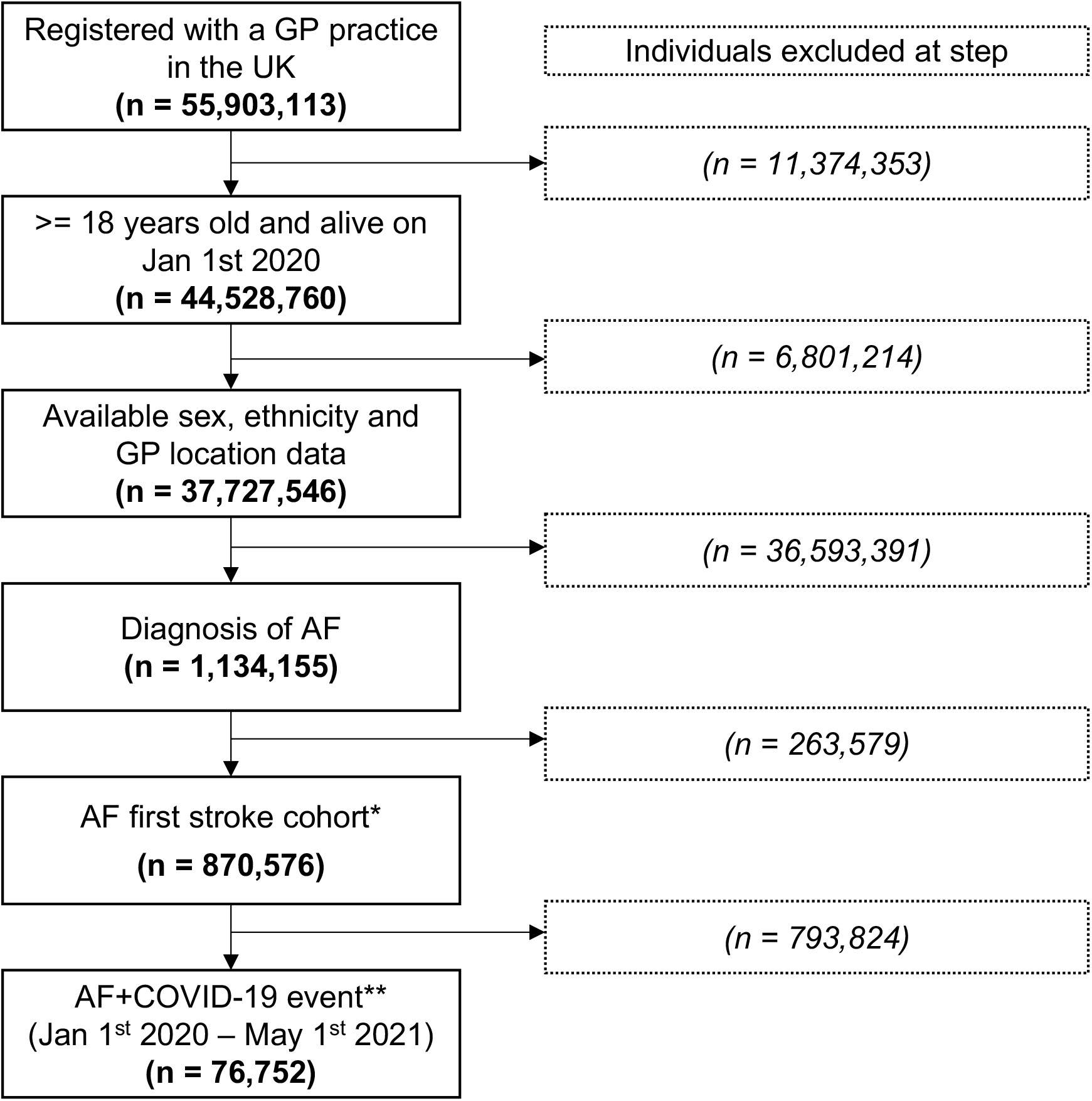
cohort inclusion flowchart showing the number of individuals excluded at each step *Excludes individuals with <5 recorded medical codes, any type of stroke diagnosis prior to AF diagnosis or within two months of AF diagnosis or after May 1st 2021 study end date. **Excludes individuals without a recorded positive COVID-19 event.

## SUPPLEMENTARY TABLES

**Supplementary Table 1.** summary of model performance statistics (AUCs) for predicting first ischaemic stroke after AF in sub-groups. 95% confidence intervals (CI) in brackets. Bold font marks highest performing model for each metric.

| Model | Female | Male | <65 years old | >=65 years old | White | Asian or asian british | Black or black british | Mixed ethnicity | Other ethnic group |
| --- | --- | --- | --- | --- | --- | --- | --- | --- | --- |
| **CHA<sub>2</sub>DS<sub>2</sub>-VASc ≥2** | 0.51<br>(0.49-0.53) | 0.53<br>(0.51-0.55) | 0.50<br>(0.44-0.56) | 0.51<br>(0.49-0.53) | 0.52<br>(0.52-0.52) | 0.60<br>(0.44-0.76) | **0.43**<br>**(0.00-1.00)** | 0.19<br>(0.00-0.64) | 0.20<br>(0.12-0.28) |
| **Logistic Regression** | 0.59<br>(0.57-0.61) | 0.58<br>(0.54-0.62) | 0.58<br>(0.56-0.60) | 0.58<br>(0.54-0.62) | 0.58<br>(0.56-0.60) | 0.54<br>(0.50-0.58) | 0.24<br>(0.00-0.73) | **0.56**<br>**(0.00-1.00)** | 0.34<br>(0.10-0.58) |
| **LSTM** | 0.53<br>(0.49-0.57) | 0.53<br>(0.45-0.61) | 0.52<br>(0.48-0.56) | 0.52<br>(0.46-0.58) | 0.53<br>(0.47-0.59) | 0.55<br>(0.37-0.73) | 0.26<br>(0.00-0.85) | 0.24<br>(0.00-0.93) | **0.57**<br>**(0.35-0.79)** |
| **Random Forest** | **0.62**<br>**(0.60-0.64)** | **0.60**<br>**(0.58-0.62)** | **0.61**<br>**(0.57-0.65)** | 0.60<br>(0.58-0.62) | 0.60<br>(0.58-0.62) | **0.65**<br>**(0.41-0.89)** | 0.29<br>(0.00-0.98) | 0.34<br>(0.00-1.00) | 0.56<br>(0.01-1.00) |
| **Transformer** | 0.58<br>(0.52-0.64) | 0.57<br>(0.53-0.61) | 0.58<br>(0.52-0.64) | 0.58<br>(0.54-0.62) | 0.58<br>(0.58-0.58) | 0.51<br>(0.37-0.65) | 0.32<br>(0.00-1.00) | 0.52<br>(0.00-1.00) | 0.34<br>(0.16-0.52) |
| **XGBoost** | 0.61<br>(0.55-0.67) | **0.60**<br>**(0.58-0.62)** | 0.59<br>(0.57-0.61) | **0.61**<br>**(0.57-0.65)** | **0.61**<br>**(0.59-0.63)** | 0.59<br>(0.34-0.84) | 0.30<br>(0.00-0.93) | 0.33<br>(0.00-1.00) | 0.49<br>(0.00-1.00) |

**Supplementary Table 2.**
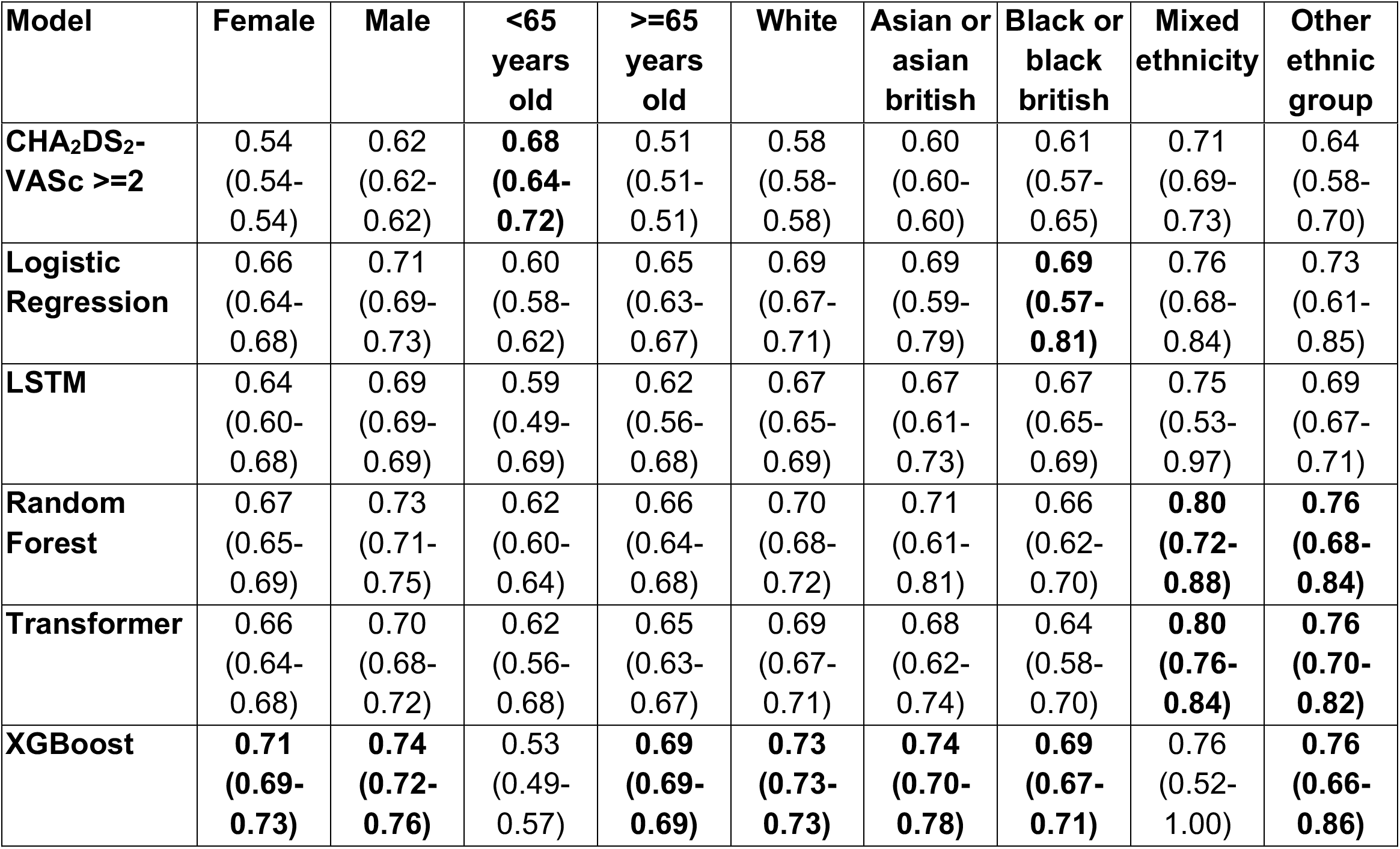
summary of model performance statistics (AUCs) for predicting COVID-19 death after AF in sub-groups. 95% confidence intervals (CI) in brackets. Bold font marks highest performing model for each metric.

| Model | Female | Male | <65 years old | ≥65 years old | White | Asian or asian british | Black or black british | Mixed ethnicity | Other ethnic group |
| --- | --- | --- | --- | --- | --- | --- | --- | --- | --- |
| **CHA<sub>2</sub>DS<sub>2</sub>-**<br>**VASc ≥2** | 0.54<br>(0.54-0.54) | 0.62<br>(0.62-0.62) | **0.68**<br>**(0.64-**<br>**0.72)** | 0.51<br>(0.51-0.51) | 0.58<br>(0.58-0.58) | 0.60<br>(0.60-0.60) | 0.61<br>(0.57-0.65) | 0.71<br>(0.69-0.73) | 0.64<br>(0.58-0.70) |
| **Logistic**<br>**Regression** | 0.66<br>(0.64-0.68) | 0.71<br>(0.69-0.73) | 0.60<br>(0.58-0.62) | 0.65<br>(0.63-0.67) | 0.69<br>(0.67-0.71) | 0.69<br>(0.59-0.79) | **0.69**<br>**(0.57-**<br>**0.81)** | 0.76<br>(0.68-0.84) | 0.73<br>(0.61-0.85) |
| **LSTM** | 0.64<br>(0.60-0.68) | 0.69<br>(0.69-0.69) | 0.59<br>(0.49-0.69) | 0.62<br>(0.56-0.68) | 0.67<br>(0.65-0.69) | 0.67<br>(0.61-0.73) | 0.67<br>(0.65-0.69) | 0.75<br>(0.53-0.97) | 0.69<br>(0.67-0.71) |
| **Random**<br>**Forest** | 0.67<br>(0.65-0.69) | 0.73<br>(0.71-0.75) | 0.62<br>(0.60-0.64) | 0.66<br>(0.64-0.68) | 0.70<br>(0.68-0.72) | 0.71<br>(0.61-0.81) | 0.66<br>(0.62-0.70) | **0.80**<br>**(0.72-**<br>**0.88)** | **0.76**<br>**(0.68-**<br>**0.84)** |
| **Transformer** | 0.66<br>(0.64-0.68) | 0.70<br>(0.68-0.72) | 0.62<br>(0.56-0.68) | 0.65<br>(0.63-0.67) | 0.69<br>(0.67-0.71) | 0.68<br>(0.62-0.74) | 0.64<br>(0.58-0.70) | **0.80**<br>**(0.76-**<br>**0.84)** | **0.76**<br>**(0.70-**<br>**0.82)** |
| **XGBoost** | **0.71**<br>**(0.69-**<br>**0.73)** | **0.74**<br>**(0.72-**<br>**0.76)** | 0.53<br>(0.49-0.57) | **0.69**<br>**(0.69-**<br>**0.69)** | **0.73**<br>**(0.73-**<br>**0.73)** | **0.74**<br>**(0.70-**<br>**0.78)** | **0.69**<br>**(0.67-**<br>**0.71)** | 0.76<br>(0.52-1.00) | **0.76**<br>**(0.66-**<br>**0.86)** |

## Notes

### Competing Interest Statement

The authors have declared no competing interest.

### Author Declarations

The data used in this study are available in NHS Digital's TRE for England, but as restrictions apply they are not publicly available (https://digital.nhs.uk/coronavirus/coronavirus-data-services-updates/trusted-research-environment-service-for-england). The CVD-COVID-UK/COVID-IMPACT programme led by the BHF Data Science Centre (https://www.hdruk.ac.uk/helping-with-health-data/bhf-data-science-centre/) received approval to access data in NHS Digital's TRE for England from the Independent Group Advising on the Release of Data (IGARD) (https://digital.nhs.uk/about-nhs-digital/corporate-information-and-documents/independent-group-advising-on-the-release-of-data) via an application made in the Data Access Request Service (DARS) Online system (ref. DARS-NIC-381078-Y9C5K) (https://digital.nhs.uk/services/data-access-request-service-dars/dars-products-and-services). The CVD-COVID-UK/COVID-IMPACT Approvals & Oversight Board (https://www.hdruk.ac.uk/projects/cvd-covid-uk-project/) subsequently granted approval to this project to access the data within the TRE for England. The de-identified data used in this study were made available to accredited researchers only. Analyses were conducted by approved researcher (AH) via secure remote access to the TRE. Only summarised, aggregate results were exported, following manual review by the NHS Digital 'safe outputs' escrow service, to ensure no output placed in the public domain contains information that may be used to identify an individual17. The North East-Newcastle and North Tyneside 2 research ethics committee provided ethical approval for the CVD-COVID-UK/COVID-IMPACT research programme (REC No 20/NE/0161).

